# Family Leaders Communicate Risk During Cascade Screening after Sudden Cardiac Death in the Young

**DOI:** 10.1101/2024.02.01.24302009

**Authors:** Lisa M. Dellefave-Castillo, Franceska Hinkamp, Lisa Shah, Courtney L. Scherr, Jennifer Young, Gregory Webster, Debra Duquette

**Author notes:** **CORRESPONDANCE:** Lisa Dellefave-Castillo, MS, CGC Center for Genetic Medicine, 303 E. Superior Street, SQ 5-408, Chicago, IL 60611. Equal contributions (Gregory Webster, Debra Duquette).

## Abstract

**Introduction:** Relatives of a victim of sudden cardiac death in the young (SCDY) may be at risk for hereditary cardiomyopathies and arrhythmias. Family leaders are often responsible for communicating risk to surviving family at a difficult time.

**Purpose:** Explore barriers and facilitators to communication about cascade screening in families who have lost a family member to SCDY

**Methods:** Semi-structured interviews (n = 14) were conducted with family members of a SCDY decedent. Participants were recruited from the Sudden Arrhythmia Death Syndrome advocacy group. Interviews were conducted until data saturation was reached. Interviews were audio recorded, transcribed, and analyzed using conventional content analysis.

**Results:** Five categories were identified from the interviews: 1. Participants understood fundamental risks but the clinical variability in arrhythmia and cardiomyopathy was difficult to interpret and convey; 2. Family leaders felt some family disregarded risk information; 3. Grief interfered with communication; 4. Communication aids were insufficient stand-alone interventions; 5. Families advocated for a “genetic family navigator”.

**Conclusion:** The five categories provide practical strategies to improve clinical care and communication for families after a SCDY and emphasize the need for genetic family navigators to facilitate cascade screening.

**GRAPHICAL ABSTRACT:** 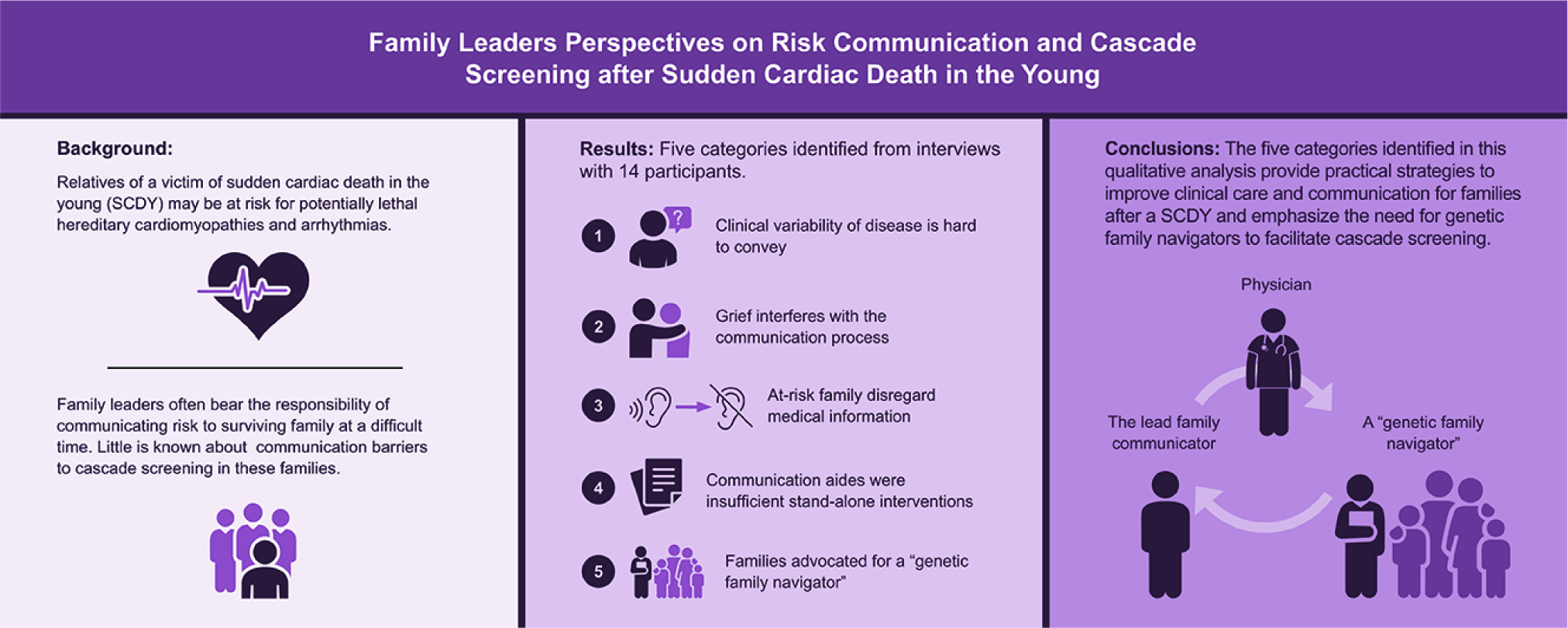

## INTRODUCTION

In people under age 45, hereditary cardiomyopathies and arrhythmias are common causes of sudden cardiac death in the young (SCDY).(1) Oftentimes, these conditions remain undetected until the SCDY event.(2,3) These conditions are typically inherited in an autosomal dominant pattern, conferring a 50% inheritance risk to all first-degree relatives with incomplete penetrance and variable expressivity.(4–6) Expert consensus statements from the American Heart Association, the American College of Cardiology, the Heart Rhythm Society, and the European Society of Cardiology recommend clinical assessment (including genetic testing) of the decedent and first-degree relatives of SCDY decedents to identify a probable cause of death and guide potentially preventative treatment when indicated.(7–10). This systematic approach of identifying and screening relatives at risk is known as cascade screening. Genetic tests for variants in cardiac rhythm disease reveal pathogenic or likely pathogenic variants in 20-30% of SCDY decedents.(11,12) When one of these variants is discovered, and cascade screening is undertaken, the average yield of cascade screening has been 9 pre-symptomatic gene-positive individuals per family.(11,13) Surviving relatives of SCD victims have low rates of sudden death when guideline-based care is applied.(14)

Despite recommendations and studies demonstrating improved outcomes, less than half of relatives are informed of their risk and even fewer receive recommended genetic testing.(15–17) Additionally, there is no standardized best practice to effectively disseminate cascade screening recommendations.(18,19) Due to policies in the United States aimed to protect the privacy of individual health information such as the Health Insurance Portability and Accountability Act (HIPAA), clinicians can help identify at risk relatives from the pedigree and encourage their patients to share risk information with their family, but they often do not initiate these conversations directly with relatives.(20,21) Some tools have been developed to provide support for this risk communication process including clinician letters, downloadable family contact kits, and private web-based family communication platforms. Even with these tools, family risk communication and cascade screening continue to be a complex and challenging process for families and their healthcare providers, especially after a SCDY event.

In this study, we sought to elicit the experiences of key family leaders regarding the cascade screening process after SCDY, focusing on several stages of the risk communication process. Our goal was to identify successes and challenges in risk communication in order to inform better family communication strategies.

## MATERIALS AND METHODS

### Participants and Recruitment

Participants were self-identified family leaders who were recruited from the distribution list of the Sudden Arrhythmia Death Syndrome (SADS) Foundation, a US-based non-profit organization that provides support to families of children and adults who are genetically predisposed to sudden death due to heart rhythm abnormalities. Inclusion criteria were age > 18 years, English-speaking, and a family member of an SCDY decedent between 1 and 39 years old. The participant needed to self-identify as a lead communicator of risk information to their family and to have contact with at least 2 other family members. The death had to occur more than one year prior to the interview to allow time for the initiation of family risk communication.

### Data Collection Tools and Procedure

A semi structured interview was used to collect participant demographics and experiences related to family risk communication after SCDY. The interview guide was created by the multidisciplinary research team and piloted with two adults who met eligibility criteria whose feedback was incorporated (development responses were not included in the data set). The specific questions and detailed methods for question development are included in the **Supplemental Methods**. A portion of the interview guide was dedicated to discussing actual communication aids that have been used in the clinical setting, which included the Dear Family Letter, SADS Pedigree Toolkit and Kintalk.org.(22,23) “Dear Family Letter” templates remain available and kintalk.org remains an active resource at the time of publication, but the SADS Pedigree ToolKit is no longer publicly available. Interviews were conducted by phone (author FH). Participants were offered a $25 gift card in appreciation for their time.

### Data Analysis

Interviews were audio recorded, transcribed verbatim, and deidentified. Conventional content analysis was used for data analysis(22). Detailed analysis methods are provided in **Supplemental Methods**. Data saturation was achieved at the 12^th^ interview.

## RESULTS

### Participants

Interviews were conducted with 14 self-identified family leaders (85% female, median age 53.5 years, (**Table 1**). Nine (64%) were parents of 1 or more decedents, 4 (29%) were siblings, and 1 (7%) was a child of a decedent (**Table 2**). One participant (7%) had lost four family members to sudden cardiac death and another three participants (21%) had lost two family members. Annual median self-reported annual household income of participants was $120,000 and most participants reported active health insurance at the time of this study (*n* = 12; 86%); two (14%) were uninsured.

**Table 1.** Participant Demographics.

| *P* | *Gender* | *Age* |
| --- | --- | --- |
| 1 | Female | 30s |
| 2 | Female | 50s |
| 3 | Female | 40s |
| 4 | Female | 40s |
| 5 | Female | 70s |
| 6 | Female | 50s |
| 7 | Female | 40s |
| 8 | Female | 50s |
| 9 | Female | 50s |
| 10 | Male | 50s |
| 11 | Female | 70s |
| 12 | Female | 40s |
| 13 | Female | 40s |
| 14 | Male | 60s |

P= participant identifier number

**Table 2.** Family communication contextual factors.

| ***P*** | ***Decedent Relationship (age in years)*** | ***Diagnosis in Decedent*** | ***Event that Initiated Cascade Screening in the Family*** | ***# of Family Members Contacted*** | ***Strategies or Tools Offered in the Communication Process*** | ***GC Engagement in Cascade Process*** |
| --- | --- | --- | --- | --- | --- | --- |
| **1** | Sibling (teen) | SUDS | Participant dx (LQTS) | 18 | None | Delayed |
| **2** | I- Child (teen)<br>II-Child (20s) | I- SUDS<br>II- LQTS* | Death of child (Decedent II) | 20 | SADS pedigree toolkit | Delayed |
| **3** | I- Sibling (20s)<br>II- 2<sup>nd</sup> (teen) | I- SUDS<br>II- SUDS | Participant dx (LQTS) | 11 | None | Delayed |
| **4** | Child (20s) | SUDS | Decedent death | 22 | SADS pedigree toolkit | Delayed |
| **5** | I- Sibling (teen)<br>II- Sibling (teen)<br>III- Sibling (20s)<br>IV- 2<sup>nd</sup> (teen) | I- SUDS<br>II- SUDS<br>III- SUDS<br>IV- LQTS | Participant dx (LQTS) | 8 | Research team | Delayed |
| **6** | Child (teen) | LQTS* | Decedent death | 35 | Research team | None |
| **7** | Child (teen) | SUDS | Decedent death | 40 | Genetic Counselor | Immediate |
| **8** | Child (<10) | SUDS | Family member dx (LQTS) | 8 | None | None |
| **9** | Child (teen) | LQTS | Decedent death | 30 | None | None |
| **10** | Child (teen) | SUDS | Decedent death | 50 | None | None |
| **11** | I- Sibling (20s)<br>II- Sibling (<10) | SUDS | Family member dx (LQT) | 25 | Physician | Delayed |
| **12** | Child (teen) | SUDS | Decedent death | 50 | None | None |
| **13** | Parent (30s) | SUDS | Participant dx (cardiomyopathy) | 6 | None | None |
| **14** | Child (teen) | LQTS | Decedent death | 35 | Research team | None |

The “cascade initiating event” was the event that triggered pedigree-based clinical evaluations. Decedent relationship is interpreted as the blank in “The decedent was the participant’s \_\_\_\_\_”.
P= participant number; dx=diagnosis; I/II/III/IV = the order in which SCDY decedents passed away if there were multiple in a family; LQTS = Long QT Syndrome; CPVT= Catecholaminergic Polymorphic Ventricular Tachycardia; GC= genetic counselor; SADS = Sudden Arrhythmia Death Syndrome Foundation; \* = diagnosed by genetic autopsy; 2<sup>nd</sup> refers to a second-degree relative, in this study either a niece or nephew.

### Decedents

Across the 14 families, there were 20 decedents with a median age of 18 years. Autopsy was performed in most cases (*n* = 14; 70%) and relatively few (*n* = 6; 30%) received post-mortem genetic testing. The cause of death was unknown in 64% of decendents. For the 9 decedents whose cause of death was confirmed, the cause was determined by molecular diagnosis, a post-mortem review of records, or a clinical diagnosis before their death. Long QT Syndrome was the predominant diagnosis made in this cohort.

For half of participants, the risk communication process began more than 15 years prior to this study. The shortest interval from initation of risk communication to its conclusion in the family was 2 years. The longest reported elapsed time was 42 years. The median number of family members contacted repoted by participants was 23.5 (range = 6-50).

### Qualitative Results

Five categories were identified from the interviews: 1. Participants understood fundamental risks but the clinical variability in arrhythmia and cardiomyopathy was difficult to interpret and convey; 2. Family leaders felt some family disregarded risk information; 3. Grief overwhelmed the communication process; 4. Communication aids were insufficient stand-alone interventions; 5. Families advocated for a “genetic family navigator”.

#### Participants understood fundamental risks to family, but clinical variability of disease was difficult to convey

Initiation of risk communication within the family and subsequent cascade screening varied across participants. Risk communication began immediatedly after SCDY for half of the participants, before they even had a diagnosis in the family or decedent. The other half did not start talking about risk until after they had a confirmed diagnosis of a genetic cardiac disease. Family leaders recalled receiving information about inherited cardiac risks from multiple professionals - predominantly cardiologists or electrophysiologists, and less frequently from ICU nurses, genetic counselors, or pediatricians. Many participants recalled receiving family risk information on more than one occurrence through these sources.

Most participants (*n*=11; 79%) recalled that cardiac risks related to SCDY could be genetic and that the cardiac condition could be dominantly inherited. Most also recalled receiving counselling that immediate family members of the decedent should undergo screening (*n*=12; 86%).

> *“I think the minute I had it and I realized I had inherited what my [parent] had, I was like I have a responsibility to share that with my cousins.” [P13]*

Several participants recalled being told about risks to family members and being instructed to contact those family members regarding possible cardiac risks, but they explained that they received little guidance for identifying at risk relatives. Furthermore, they felt ill-equiped to navigate the process of communicating risk to family members themselves, had little structure for organizing clinical referrals in the family, and often were struggling to work through their own emotions.

Participants shared that the clinical variability of a suspected or confirmed genetic cause of SCDY was challenging to understand. Even at the time of our interviews, families identified the complex interplay between variable expression and reduced penetrance as an ongoing barrier to understanding and thus preventing effective communication.

> *“ I mean, it wasn’t until* further *research that I realized the severity of the condition was not necessarily transferred at a, um, like equal rate. It wasn’t like - Um, I guess that was how it presented in each case. Like how consistent would the presentation be across the family.” [P1]*

#### Family disregarded risk information

Many family leaders reported non-cooperation from family. They expressed frustration with family members who knew about their potential risks but disregarded the information by verbally denying it or not completing the recommended screening. Participants frequently explained relatives’ denial and inaction as a response to fear of their cardiac risk, a difference in risk tolerance, a difference in risk perception, or fatalism, “If it happens; it happens”. In some cases, family members doubted the accuracy or validity of information because of the family leader’s lack of medical credibility.

> *“I think it would have been good for the medical people that we dealt with to set up a visit with my entire family, my siblings, the parents to explain what they thought caused it and how close to their family this could impact them* and *I think that would have went over a lot better than me telling them.”[P10]*

Interestingly, two participants were medical professionals, and both reported that their expertise was discounted within the family.

#### Grief overwhelmed the communication process

Family leaders said that the emotional weight of a SCDY hindered effective communication in three critical tasks: (1) understanding risk information; (2) comprehending the possibilty of a personal diagnosis as a surviving relative; and (3) coherently communicating information while grieving. First, several participants said that they heard the report about the decedent, but could not really process it.

> *“I’ll be honest, I wasn’t in the frame of* mind *that I could have successfully parsed the information, anyway. Well, my [child] was my best friend… then I lost [them]. I broke. I broke pretty badly.” [P8]*

The shock produced by the SCDY diagnosis was described as a barrier to understanding the risk information, especially in those who subsequently received a personal cardiac diagnosis which clarified the cause of the decedent’s death.

> *“I think the shock of being like, ‘Holy crap, I just figured out how my [sibling] died,’ was just bouncing around in my head and so to really focus on what was happening and the gravity of it all was just a lot to take in for that one day.” [P3]*

Most family leaders said that their grief made it difficult to effectively communicate risk information.

> *“I think the hardest part for me is just it being the reality of talking about it… at the end of the day, it’s hard emotionally to talk about… I think grief gets in the way, I think that honestly is kind of the bottom line for me, and some days I can* face *reality and some days I can’t.” [P4]*

Participants also described difficulty balancing ongoing obligations in their life after the SCDY such as taking care of other surviving children and “keeping them alive.”

#### Communication aids were insufficient stand-alone interventions

Family leaders emphasized the need for direct-to-family communication that included medical professionals. The degree of genetic counselor engagement in the family communication process varied with half reporting no involvement. Of the participants who had genetic counseling, only one had a genetic counselor who was involved in the cardiac risk evaluation and family screening support. Others reported delayed involvement, where they met with a genetic counselor after the family risk communication had begun.

Only half of the participants reported receiving specific tools or strategies to assist with risk communication among family members. All three communication aids discussed in the interviews were developed with input from medical experts (“Dear Family” letter, SADS pedigree toolkit, Kintalk.org). (22,23) When asked to review these aids during the interview, participants only assigned medical credibility to the “Dear Family” letter. This letter, on physician letterhead and signed by a physician, was identified as more desirable than the others because it carried the most prominent appearance of medical credibility. Participants discussed the potential of these tools to elicit a negative reaction from recipient family members or to be disregarded altogether by unreceptive family members. Six participants voiced the potential for these tools to be shocking if used as the first method to communicate risks and recommendations.

> *“For me personally, I don’t want to have to read it in a letter. I mean, if somebody sent me a letter, and someone told me, you know, someone in your family died suddenly, that would have freaked me out… I would not want that to be done to my family* members*.”*[P12]

A similar critique across communication aids was that they may be ineffective due to a lack of personalization.

> *“It doesn’t really give you an impact. The problem is it’s, to me, because it’s an emotional issue you have to hit the emotions. A letter is very* impersonal *and it tends to minimize things. You’re trying to gently word things …… The more you genericize [sic] it the less impact you’re going to have.”*[P8]

Although some participants considered aids as potentially useful, participants echoed that a single tool is insufficient as a stand-alone intervention.

> *“If I could’ve snapped my fingers and had that resource available, that would’ve been great. But there’s no pamphlet. There’s no video. There’s no book. There’s nothing that can convey it like the words, and the right words, and the right tone. So, people skilled in that would be valuable.”[P14]*

#### Families advocated for a “genetic family navigator”

A few participants had been enrolled in a genetic research protocol and they reported a highly facilitated family risk communication process. In these cases, a multidisciplinary cardiac research team identified at-risk relatives and with the help and permission of the participant, directly contacted them with information regarding genetic risks as a part of study enrollment. These participants believed more family members underwent screening because of this facilitation and because there was no cost for genetic testing or cardiac screening for the family.

Almost all family leaders suggested it would have been helpful to have a provider with genetic expertise and the capacity to navigate screening for a family. Many participants believed a genetic counselor should fill this role or described the value of a case manager. To synthesize these concepts, we employed the general term “genetic family navigator.”

Participants suggested that a genetic family navigator would be crucial at several time points. In the early stages of communication, participants suggested that a genetic family navigator could facilitate group education within the family, add credibility to the message, interpret complex technical information, provide anticipatory guidance, initiate follow up for families’ cascade process and support psychosocial needs.

> *“…just say that there was a representative that counsels family members on their risk of this occurring to them … I mean, I think that it’s essential… would’ve been very helpful is if a day or two later there was someone that had this service available and was available to relatives.”* [P9]

Family leaders reported struggling to provide the detailed medical education to family as well as providing the right tone and emphasis for that information.

> *“And that was hard too, to get this information to people to* understand*, relatives to understand that we weren’t looking for clogged arteries or bypass surgeries but this is the electrical workings of the heart versus a clog or a valve that’s too larger. So just trying to relay that information was also difficult because not everybody had a good understanding…[I] pretty much spoke to them just like I’m speaking to you now. I think the frustrating part for me were them not understanding fully the risk or how genetics work.”* [P7]

Almost half of the participants described the primary benefit of this type of model in family communication would relieve the burden for bereaved family members.

> *“I would say the hardest part was that I had to do it myself, as far as it came to really knowing what was going on with (decedent)… it would have been easier if a doctor had said, “Hey, okay, let me explain these to you, and this is what we need to do. I’ll contact your family.” No one had done that. I had to do it all myself. So, that was hard … it didn’t really give me time to grieve a whole lot.”[P12]*

The minority of participants who had substantial guidance in some or all of the stages of family communication reported that they valued the step-by-step guidance and provider availability that they experienced.

## DISCUSSION

Discussing family risk for SCDY and recommended genetic screening following an SCDY is a complex and challenging process further impacted by the grief surrounding the context. Results from our interviews demonstrated that family members struggle to communicate the complexity of genetic heart disease with their family, particularly while they were grieving the loss of their family member. They felt that existing communication aids were insufficient stand-alone interventions to facilitate risk communication and promote uptake of genetic screening, particularly among reactant family members. These experiences led participants to suggest the need for a “genetic family navigator”. Recommendations stemming from this study to address these challenges with family leaders and other at-risk family members are presented in Table 3.

**Table 3.** Summary of Participant-Based Clinical Recommendations.

| **Study findings** | **Clinical Recommendations following SCDY** |
| --- | --- |
| Use continuous and non-linear risk communication; communicate technical information carefully and repeatedly | Consider family communication as an ongoing, cyclic process. |
|  | Discuss the evolving needs of families as they receive follow up care. |
|  | Clearly explain the technical aspects of genetic risk including inheritance pattern, the identification of relatives at risk. |
|  | Counsel carefully about the extensive variability in clinical manifestations between family members. |
|  | Provide specific screening instructions for relatives at risk. |
| Some family members will disregard risk information | Anticipate that resistance among relatives at risk is likely. Offer anticipatory guidance and normalize frustration. |
| Grief overwhelmed the communication process | Acknowledge that grief may be the biggest barrier at each stage of risk communication and a barrier to cascade screening for relatives at risk. |
|  | Triage family members for long term psychological support. |
| Communication aids were insufficient stand-alone interventions | Participants preferred communication aids as a supplement to direct contact by the communicator. |
|  | Use the following elements in communication aids: personalization, emotional connection to the recipient, symbols of medical credibility. |
|  | Offer multiple strategies and tools and listen to families about what type of communication aid will be most useful |
| Families advocated for genetic family navigators | The job of “genetic family navigator” combines the current roles that include genetic counselors, case managers, and medical providers (such as doctors and nurses). |

While the challenges of family risk communication have been described in other conditions (20,24), there are barriers that are specific to SCDY. One prominent example from our interviews was that families initiated the cascade screening process at different times. In our study, risk communication sometimes began with the diagnosis of the decedent immediately post-arrest, other times the process of identifying the decedent’s phenotype was restarted posthumously, or was initiated after a living family member’s diagnosis shed light on the cause of the SCDY. Expert consensus statements recommend a standardized approach to family screening after a SCDY;(25,26) however, the decentralized coroner/medical examiner system in the United States and variable practices among clinicians introduce substantial heterogeneity into the family experience. As the family communication process is ongoing, clinicians should not assume that adequate counseling has been done based on the time since the SCDY, nor should they assume that family communication has been linear or uniform. In addition, the standardized approach recommended in current consensus documents offers a path forward to reduce variability in the future.

Participants recognized their role as a communicator of risk information to their family. They recalled learning the possible genetic cardiac risks related to the SCDY, the concept of dominant inheritance, and the justification for cardiac screening of immediate and extended family. This is important because prior work has determined that people with an understanding of autosomal dominant risk in genetic heart condition conditions were more likely to communicate with their at-risk siblings and children.(27) While participants understood this information for themselves, they had difficulty conveying concepts like variable expression and reduced penetrance. Participants retrospectively felt that it would have been helpful to have a better understanding of these concepts to allow them a better chance at successful risk communication.

Our participants expressed frustration with those family members who seemed to have a lower perception of cardiac risk if they or other family members were asymptomatic, despite a family member dying suddenly, and most notably in those who had suffered the loss of an apparently healthy, asymptomatic family member. Notably, this frustration is not necessarily the case in all cardiac diseases with significant arrhythmia risk.(28) Additionally, even years after the event, some participants described family members’ resistance to undergo clinical screening due to fear or low risk perception. Lack of interest and denial from at-risk family members and accompanying frustration is a common experience within family communication research.(28–31) Studies on the attitudes of ambivalent family members are scarce, perhaps due to the lack of willing family members to participate in research about resistance. Providing anticipatory guidance regarding family resistance and frustration may be helpful to better anticipate and lower the family members’ frustration, who share the risk information with their family.(21)

All of the participants of this study endured the tragedy of at least one family member lost to SCDY and recognized that grief and shock were significant barriers to risk communication. A unique tension exists for risk communication in the family after a SCDY. Timely communication is imperative due to cardiovascular risk but the emotional shock and grief can deadlock this family-driven dissemination process. Ingels and colleagues assessed psychological functioning in bereaved family members after a SCDY and found that approximately 50% of family members reported significant psychological difficulties such as anxiety, prolonged grief and posttraumatic stress symptoms on average 6 years after the SCDY.(32) Van der Werf and colleagues found that some first-degree relatives of SCD decedents who attended cardiogenetics evaluations postponed their evaluation due to emotional pain.(33) It is critical that clinicians recognize how emotions can supersede an individual’s ability to understand, plan and communicate SCDY risk information and that these families may need extra support in the cascade screening process.

Existing communication aids discussed with participants were collectively described as insufficient as stand-alone interventions because they were potentially shocking, lacking personalization or lacking in medical credibility. Based on the information provided by the participants and the need for personalization, a tool should contain components of familiarity and emotional connection to the recipient, and posess medical credibiliity. The logic of precision health should extend to the types of resources given to families so that it is the right intervention, for the right person, at the right time.(34)

Interestingly, while all three communication aids discussed were developed with input from medical professionals, participants only assigned medical credibility to the “Dear Family” clinic letter, which is often on physician letterhead and signed by a physician, and thus more desirable. Communication aids that are personalized, presented with a symbol of medical credibility and used as a follow up to person-to-person contact may be more effective.

Almost all participants enthusiastically endorsed the idea of the assistance of a genetic family navigator to facilitate risk communication after SCDY. This role was described as hands-on, beholden to an entire family, and acting as a coordinator, educator and communicator of risk information to at-risk relatives. Research is mixed on the role that providers versus family members should play during cascade screening, and there may be condition-specific factors that influence what facilitates this process.(31,35,36) For SCDY, direct contact by a provider with the consent of the index patient may overcome some of the barriers entrenched in communication between family members as outlined in this study. Direct contact has been proposed in previous studies to facilitate medical navigation and family communication for those with a history of inherited cardiac disease and SCDY(28,37) as well as other adult onset conditions.(31,38–41) Our study adds another data point in the cardiovascular and sudden death space where family leaders who consider themselves the risk communicators of their family would support direct relative contact from a genetic family navigator to facilitate this critical process.

## LIMITATIONS

The participants in this study were volunteers, reached through a SCDY interest group. They were likely enriched for highly motivated communicators. While our sample achieved diverse communication perspectives, there was limited demographic diversity.

## CONCLUSIONS

Family leaders identified several barriers to effective cascade screening in the family. A more active approach by a medical professional facilitator was the resounding proposed intervention. We characterized five categories of family risk communication about SCDY and documented that direct contact is welcomed by this population.

## ETHICS DECLARATION

This study received approval from the Northwestern University Institutional Review Board (STU00208325). All participants provided electronic informed consent before completing the demographic survey and interview. Transcripts from the interviews were de-identified before the analysis. This study was conducted in accordance with the Helsinki Declaration.

## Data Availability

Transcripts redacted for participant privacy are available upon request, contingent on appropriate human subjects protection and after execution of an adequate data sharing agreement.

## ACKNOWLEDGEMENTS

We would like to acknowledge all of the participants of this study who took the time to share with us some difficult information. We would also like to acknowledge the SADS Foundation who were instrumental in the recruitment of these participants. Liz Perez provided graphic design for the abstract.

## AUTHOR CONTRIBUTIONS

Conceptualization (LDC, FH, CS, LS, DD), Data curation (FH), formal analysis (LDC, FH, LS), supervision (LDC, CS, DD), writing-original draft (FH), writing-review and editing (LDC, JY, GW).

## REFERENCES

1. Wilde AA, Behr ER. Genetic testing for inherited cardiac disease. Nat Rev Cardiol 2013;10:571–583.

2. Gray B, Ackerman MJ, Semsarian C, Behr ER. Evaluation After Sudden Death in the Young: A Global Approach. Circ Arrhythm Electrophysiol 2019;12:e007453.

3. D’Ascenzi F, Valentini F, Pistoresi S et al. Causes of sudden cardiac death in young athletes and non-athletes: systematic review and meta-analysis: Sudden cardiac death in the young. Trends Cardiovasc Med 2022;32:299–308.

4. Spoonamore KG, Ware SM. Genetic testing and genetic counseling in patients with sudden death risk due to heritable arrhythmias. Heart Rhythm 2016;13:789–797.

5. Girolami F, Frisso G, Benelli M et al. Contemporary genetic testing in inherited cardiac disease: tools, ethical issues, and clinical applications. J Cardiovasc Med (Hagerstown) 2018;19:1–11.

6. Franciosi S, Abrams DJ, Ingles J, Sanatani S. Sudden Cardiac Arrest in the Paediatric Population. CJC Pediatr Congenit Heart Dis 2022;1:45–59.

7. Al-Khatib SM, Stevenson WG, Ackerman MJ et al. 2017 AHA/ACC/HRS guideline for management of patients with ventricular arrhythmias and the prevention of sudden cardiac death: Executive summary: A Report of the American College of Cardiology/American Heart Association Task Force on Clinical Practice Guidelines and the Heart Rhythm Society. Heart Rhythm 2018;15:e190–e252.

8. Priori SG, Blomström-Lundqvist C, Mazzanti A et al. 2015 ESC Guidelines for the management of patients with ventricular arrhythmias and the prevention of sudden cardiac death: The Task Force for the Management of Patients with Ventricular Arrhythmias and the Prevention of Sudden Cardiac Death of the European Society of Cardiology (ESC). Endorsed by: Association for European Paediatric and Congenital Cardiology (AEPC). Eur Heart J 2015;36:2793–2867.

9. Erickson CC, Salerno JC, Berger S et al. Sudden Death in the Young: Information for the Primary Care Provider. Pediatrics 2021;148.

10. Stiles MK, Wilde AAM, Abrams DJ et al. 2020 APHRS/HRS expert consensus statement on the investigation of decedents with sudden unexplained death and patients with sudden cardiac arrest, and of their families. Heart Rhythm 2021;18:e1–e50.

11. Bagnall RD, Weintraub RG, Ingles J et al. A Prospective Study of Sudden Cardiac Death among Children and Young Adults. N Engl J Med 2016;374:2441–2452.

12. Webster G, Puckelwartz MJ, Pesce LL et al. Genomic Autopsy of Sudden Deaths in Young Individuals. JAMA Cardiol 2021;6:1247–1256.

13. Tan HL, Hofman N, van Langen IM, van der Wal AC, Wilde AA. Sudden unexplained death: heritability and diagnostic yield of cardiological and genetic examination in surviving relatives. Circulation 2005;112:207–213.

14. Müllertz KM, Christiansen MK, Broendberg AK, Pedersen LN, Jensen HK. Outcome of clinical management in relatives of sudden cardiac death victims. Int J Cardiol 2018;262:45–50.

15. Shah LL, Daack-Hirsch S, Ersig AL, Paik A, Ahmad F, Williams J. Family Relationships Associated With Communication and Testing for Inherited Cardiac Conditions. West J Nurs Res 2019;41:1576–1601.

16. Shah LL, Daack-Hirsch S. Family Communication About Genetic Risk of Hereditary Cardiomyopathies and Arrhythmias: an Integrative Review. J Genet Couns 2018;27:1022–1039.

17. Ajufo E, deGoma EM, Raper A, Yu KD, Cuchel M, Rader DJ. A randomized controlled trial of genetic testing and cascade screening in familial hypercholesterolemia. Genet Med 2021;23:1697–1704.

18. Roberts MC, Dotson WD, DeVore CS et al. Delivery Of Cascade Screening For Hereditary Conditions: A Scoping Review Of The Literature. Health Aff (Millwood) 2018;37:801–808.

19. Sturm AC. Cardiovascular Cascade Genetic Testing: Exploring the Role of Direct Contact and Technology. Front Cardiovasc Med 2016;3:11.

20. Wiseman M, Dancyger C, Michie S. Communicating genetic risk information within families: a review. Fam Cancer 2010;9:691–703.

21. Young AL, Butow PN, Tucker KM, Wakefield CE, Healey E, Williams R. Challenges and strategies proposed by genetic health professionals to assist with family communication. Eur J Hum Genet 2019;27:1630–1638.

22. Kintalk. Version 3.3.6.2.8.5 ed. University of California, San Francisco.

23. Dheensa S, Lucassen A, Fenwick A. Limitations and Pitfalls of Using Family Letters to Communicate Genetic Risk: a Qualitative Study with Patients and Healthcare Professionals. J Genet Couns 2018;27:689–701.

24. Smart A. Impediments to DNA testing and cascade screening for hypertrophic cardiomyopathy and Long QT syndrome: a qualitative study of patient experiences. J Genet Couns 2010;19:630–639.

25. Stiles MK, Wilde AAM, Abrams DJ et al. 2020 APHRS/HRS expert consensus statement on the investigation of decedents with sudden unexplained death and patients with sudden cardiac arrest, and of their families. Heart Rhythm 2021;18:e1–e50.

26. Fellmann F, van El CG, Charron P et al. European recommendations integrating genetic testing into multidisciplinary management of sudden cardiac death. Eur J Hum Genet 2019;27:1763–1773.

27. Batte B, Sheldon JP, Arscott P et al. Family communication in a population at risk for hypertrophic cardiomyopathy. J Genet Couns 2015;24:336–348.

28. Ormondroyd E, Oates S, Parker M, Blair E, Watkins H. Pre-symptomatic genetic testing for inherited cardiac conditions: a qualitative exploration of psychosocial and ethical implications. Eur J Hum Genet 2014;22:88–93.

29. Vavolizza RD, Kalia I, Erskine Aaron K et al. Disclosing Genetic Information to Family Members About Inherited Cardiac Arrhythmias: An Obligation or a Choice? J Genet Couns 2015;24:608–615.

30. Whyte S, Green A, McAllister M, Shipman H. Family Communication in Inherited Cardiovascular Conditions in Ireland. J Genet Couns 2016;25:1317–1326.

31. Hardcastle SJ, Legge E, Laundy CS et al. Patients’ perceptions and experiences of familial hypercholesterolemia, cascade genetic screening and treatment. Int J Behav Med 2015;22:92–100.

32. Ingles J, Spinks C, Yeates L, McGeechan K, Kasparian N, Semsarian C. Posttraumatic Stress and Prolonged Grief After the Sudden Cardiac Death of a Young Relative. JAMA Intern Med 2016;176:402–405.

33. van der Werf C, Onderwater AT, van Langen IM, Smets EM. Experiences, considerations and emotions relating to cardiogenetic evaluation in relatives of young sudden cardiac death victims. Eur J Hum Genet 2014;22:192–196.

34. Khoury MJ, Iademarco MF, Riley WT. Precision Public Health for the Era of Precision Medicine. Am J Prev Med 2016;50:398–401.

35. Frey MK, Ahsan MD, Bergeron H et al. Cascade Testing for Hereditary Cancer Syndromes: Should We Move Toward Direct Relative Contact? A Systematic Review and Meta-Analysis. J Clin Oncol 2022;40:4129–4143.

36. Rosén A, Krajc M, Ehrencrona H, Bajalica-Lagercrantz S. Public attitudes challenge clinical practice on genetic risk disclosure in favour of healthcare-provided direct dissemination to relatives. Eur J Hum Genet 2023.

37. Wisten A, Zingmark K. Supportive needs of parents confronted with sudden cardiac death--a qualitative study. Resuscitation 2007;74:68–74.

38. Aktan-Collan K, Haukkala A, Pylvänäinen K et al. Direct contact in inviting high-risk members of hereditary colon cancer families to genetic counselling and DNA testing. J Med Genet 2007;44:732–738.

39. Chivers Seymour K, Addington-Hall J, Lucassen AM, Foster CL. What facilitates or impedes family communication following genetic testing for cancer risk? A systematic review and meta-synthesis of primary qualitative research. J Genet Couns 2010;19:330–342.

40. Rahm AK, Sukhanova A, Ellis J, Mouchawar J. Increasing utilization of cancer genetic counseling services using a patient navigator model. J Genet Couns 2007;16:171–177.

41. Henrikson NB, Blasi P, Figueroa Gray M et al. Patient and Family Preferences on Health System-Led Direct Contact for Cascade Screening. J Pers Med 2021;11.

