## Supplemental Files for "Family Leaders Communicate Risk During Cascade Screening after Sudden Cardiac Death in the Young"

### Supplementary Materials for

#### Perspectives on Risk Communication from Family Leaders who Coordinated Cascade Screening after Sudden Cardiac Death in the Young

**AUTHORS:** Lisa M. Dellefave-Castillo, MS, CGC; Franceska Hinkamp, MS, CGC; Lisa Shah, PhD, RN; Courtney L. Scherr, PhD; Jennifer Young, PhD; Gregory Webster, MD, MPH; Debra Duquette, MS, CGC

**This PDF file includes:**

Supplemental Methods
Supplemental Table 1 (Semi-structured interview questions)

**SUPPLEMENTAL METHODS**

One-on-one interviews were conducted over the phone by FH and recorded and lasted approximately 1 hour using the semi-structured interview guide (Supplement: Semi-structured interview questions). The participants were first asked background questions related to circumstances of the death of their relative (the "decedent"). To characterize the risk communication after the sudden cardiac death in the young (SCDY) in their family, questions were designed to explore the family disclosure context. These questions provided context specific to the experiences, challenges and proposed interventions related to participants initially-receiving cardiac risk information ("receiving"), the identification of at-risk relatives and the strategizing of contacting them ("strategizing"), the actual communication of risk ("communicating"), and recommendations to at risk family members ("promoting screening"). Questions were open-ended except for basic demographics.

Interviews were transcribed verbatim and deidentified. Analysis began with each coder (FH and LS) reading the transcripts in their entireity and creating a summary of first impressions. Next, FH and LS coded two transcripts each, coding text relevant to each of the research aims. Coded text was analyzed and intitial codes were developed from the text data. Coded transcripts were sent to the other coder who reviewed coding, added codes, and noted discrepancies which were discussed. This step resulted in the initial draft of a codebook with codes and definitions. In the next round of coding, four more transcripts were reviewed using the initial codebook. In this round of coding, the codebook was further developed by adding codes and revising definitions based on the new transcripts reviewed. Again, analyst triangulation was used, and coded transcripts and revised codebook were exchanged between coders, who reviewed all coding and noted discrepencies. Discrepancies in coding were discussed and resolved by reexamining the context of the quote within the transcript and cross-referencing the original definitions assigned to the codes. Codebook creation was an iterative process in which codes and defintions were continuously refined as additional transcripts were reviewed. No further changes were made to the codebook after the 12th transcript, and we felt confident that we had reached data saturation. We reviewed the final codebook and grouped the codes into meaningful categories named by direct quotes from the data or notes from the summaries of our first impressions of the data.

#### SUPPLEMENT: SEMI-STRUCTURED INTERVIEW QUESTIONS

| **PART 1: DECEDENT BACKGROUND** | | |
|---|---|---|
| **Q#1: I would like to begin by asking you to tell me about your family member(s) who passed away.**<br>● Could you tell me their name?<br>● How old were they when they passed?<br>● How did you find out that their death(s) was a sudden death event?<br>● Did they ever receive a diagnosis related to their death?<br><br>→ Thank them for response (adopt name of family member if offered) | | |
| **PART 2: FAMILY DISCLOSURE CONTEXT** | | |
| **Investigator statement: After a sudden death event, medical professionals may recommend that blood relatives be contacted because of the possibility that they could have related health risks. They may make recommendations for what health screening tests should be ordered for at risk family members. As we move through the rest of the interview, I'd like you to think about the way that information about health risks to relatives was given to you and how you passed along that information.** | | |
| **MAIN QUESTION** | **PROBE QUESTIONS** | **CLARIFYING QUESTIONS/PIVOT STATEMENTS** |
| **Q#2: After [decedent] died, what were you told by professionals about the cardiac risks to other family members?** | Who explained this information?<br>Was a genetic counselor involved in your care?<br>What did he/she/they say the risks were? | Can you tell me more about what you mean by that?<br><br>(Professionals means health care providers, medical examiners, or coroners) |
| **Q#3: Was the information about family risk understandable?** | What, if anything was difficult to understand? | Why was this difficult to understand?<br>Can you tell me why you think that? |
| **Q#4: How might the communication of your family's cardiac risk information been improved?** | Do you think the content of information that was given to you could have been better?<br>Do you think the way the information was given could have been better?<br>How do you think things would be different if this improvement had been made? | Can you tell me more about why this would be helpful? |

(continued on next page)

| **MAIN QUESTION** | **PROBE QUESTIONS** | **CLARIFYING QUESTIONS/PIVOT STATEMENTS** |
|---|---|---|
| **Q#5: During what part of the process did you realize that you were the person responsible for telling family members about their cardiac risk?** | Why do you think you were the person to spread this information? | Can you tell me more about what you mean by that? |
| **Q#6: How did you figure out who to contact in the family about their risk?** | Did you have a system for notifying family?<br><br>Were there relatives who were at risk who you chose not to contact (Why?)<br><br>Were you provided any tools to help you organize the process of notifying and gathering information from your relatives? | Yes→ Where did this tool come from?<br>Was this tool helpful?<br><br>No→ do you think a tool should have been provided? |
| **Q#7: Can you think of anything that could have helped you to identify at risk family members?** | What about this would be helpful?<br><br>How would this have changed the way you identified at risk family members? | |
| **Q#8: What was your experience like telling family members about their cardiac risk?** | How did they react?<br><br>How did you feel when giving this information? | **Pivot: That's interesting to know. What I would like to know more about is...* |
| **Q#9: In times where you were successful in communicating cardiac risk information to your family members, what factors allowed you to be successful?** | | Can you tell me more about why you think this is? |
| **Q#10: What was challenging about communication of the cardiac risk information to family members?** | What could have gone <u>better</u> with regard to communicating about their cardiac risks? | Can you tell me more about what you mean by that? |
| **Q#11: Is there a resource that you think would have helped overcome the challenge of [stated challenge]?** | In a perfect world, what would have helped you communicate this cardiac risk information considering your [stated challenge].<br><br>How would this additional resource have helped? | |

| PART 3: CURRENT AND PROPOSED COMMUNICATION INTERVENTIONS | | |
|---|---|---|
| **There are some tools that are currently used to help families with the process of health risk communication. For the next set of questions, I would like to discuss 3 examples and get your reactions to them based on your experience. I will read a short description of each and then ask a couple of questions about your reactions.**<br>**1) Family Letter:** In some clinics, families are given a letter that says "Dear Family Member, a member of your family recently died suddenly from a cardiac condition and you may be at risk" and may provide information about health risks and/or recommended cardiac screening. If relatives want to be examined, they are encouraged to contact a genetics clinic or a cardiologist.<br>**Q#12: Do you think this would have been helpful to you? Why or why not?**<br>**Q#13 Is there a disadvantage to this tool from your perspective?**<br>**2) SADS Pedigree Kit:** This kit provides tools to help families organize the process of notifying and gathering information from relatives. The 3-step kit involves 1) filling out the "family members to contact" list by entering the names of relatives that can be used as a guide to contact 2) completing a questionnaire about symptoms, diagnosis and screening results of sudden arrhythmia death syndromes in the family and 3) contacting other family members using a template family notification letter that can be sent via mail or email.<br>**Q#14: Do you think this would have been helpful to you? Why or why not?**<br>**Q#15 Is there a disadvantage to this tool from your perspective?**<br>**3) KinTalk:** Kintalk is a social media platform which allows people with hereditary cancer syndromes to communicate with their families about genetic risk information (for this tool, imagine something similar but for cardiology). In Kintalk, a person can create a profile to and upload genetic testing results to a secure portal. They can also upload a family tree or other documentation of screening recommendations given to them. Patients can then privately invite relatives to see the information. Kintalk provides templates for messages for contacting family members to alert them about their risk level and explanations about how they may be at risk. Kintalk also has information about cancer syndromes including news about recent research, videos and downloadable pamphlets.<br>**Q#16: Do you think this would have been helpful to you? Why or why not?**<br>**Q#17: Is there a disadvantage to this tool from your perspective?** | | |
| **MAIN QUESTION** | **PROBE QUESTIONS** | **CLARIFYING QUESTIONS/PIVOT STATEMENTS** |
| **Q#18: Do you feel new resources should be developed to facilitate better communication about cardiac risks to family members?** | Can you think of any missed opportunities for tools or professional support throughout this process of communicating family risk? | |
| **Q#19: What new resources would you like to see developed?** | How would this resource have improved the process in your family? | |

| PART 4: DEMOGRAPHICS |
| --- |
| **I wanted to end this interview with some quick questions about you and your family (you are not obligated to answer but will help me understand the background of your family)**<br>❑ Age<br>❑ Ethnicity<br>❑ Gender<br>❑ Highest level education completed<br>❑ Marital Status<br>❑ Number of children<br>❑ Occupation<br>❑ Annual Income<br>❑ Health Insurance status<br>❑ How were you related to (the decedent(s))?<br>❑ Approximately how many people were contacted in the family about their health risks?<br>❑ How many were first degree relatives (ex. parents, siblings, children)<br>❑ How many were second degree relatives (ex. Half Sibling, Grandparent, Aunt, Uncle, Niece, Nephew)<br>❑ Did (the decedent(s)) have an autopsy or genetic testing which confirmed the cardiac condition?<br>❑ Did any survivors in your family undergo an echocardiogram after the death? If so, how many?<br>❑ Did any survivors in your family undergo an electrocardiogram after the death?<br>❑ Was other testing performed on the heart for any of the survivors after the death?<br>❑ Did anyone in the family other than the decedent undergo genetic testing?<br>❑ Was a disease diagnosed in anyone else in the family?<br>→ Was it the same disease that was suspected in the decedent? |
| **PART 5: FINAL THOUGHTS** |
| **Q#20: Is there anything else that you would like to share about communicating about cardiac risk in your family that I did not think to ask?**<br>[Pause] |
